# Serological Response to BNT162b2 and ChAdOx1 nCoV-19 Vaccines in Patients with Inflammatory Bowel Disease on Biologic Therapies; A Multi-Center Prospective Study

**DOI:** 10.1101/2021.10.31.21265718

**Authors:** Mohammad Shehab, Fatema Alrashed, Ahmad Alfadhli, Khazna AlOtaibi, Abdulla AlSahli, Hussain Mohammad, Preethi Cherian, Irina Alkhair, Thangavel Alphonse Thanaraj, Arshad Channanath, Hamad Ali, Mohamed Abu-Farha, Jehad Abubaker, Fahd Al-Mulla

**Affiliations:** Division of Gastroenterology, Department of Internal Medicine, Mubarak Alkabeer University Hospital, Kuwait University, Kuwait; Department of Pharmacy Practice, Faculty of Pharmacy, Health Sciences Center (HSC), Kuwait University, Jabriya, Kuwait; Department of Biochemistry and Molecular Biology, Dasman Diabetes Institute (DDI), Dasman, Kuwait; Department of Genetics and Bioinformatics, Dasman Diabetes Institute (DDI), Dasman, Kuwait; Department of Medical Laboratory Sciences, Faculty of Allied Health Sciences, Health Sciences Center (HSC), Kuwait University, Jabriya, Kuwait

**Keywords:** IBD, COVID19, Vaccine, immunogenicity, infliximab, adalimumab, vedolizumbab, ustekinumab

## Abstract

**Introduction:** Immunogenicity of SARS-CoV-2 vaccines in patients with inflammatory bowel disease (IBD) on biologics are not well studied. The goal of this study is to measure serological response to BNT162b2 and ChAdOx1 nCoV-19 vaccines in patients with IBD receiving different biologic therapies.

**Method:** We performed a multi-center prospective study between August 1^st^, 2021, and September 15^th^, 2021. We measured seropositivity of SARS-CoV2 antibodies, SARS-CoV-2 IgG and neutralizing antibody concentrations, in patients with IBD receiving biologic therapies between 4–10 weeks after second dose or 3-6 weeks after first dose of vaccination with BNT162b2 or ChAdOx1 nCoV-19 vaccines.

**Results:** There were 126 patients enrolled (mean age, 31 years; 60% male; 71% Crohn’s disease, 29% ulcerative colitis). 92 patients were vaccinated with BNT162b2 vaccine (73%) and 34 patients with ChAdOx1 nCoV-19 vaccine (27%). The proportion of patients who achieved positive anti-SARS-CoV-2 IgG antibody levels after receiving 2 doses of the vaccine in patients treated with infliximab and adalimumab were 44 out of 59 patients (74.5%) and 13 out of 16 patients (81.2%), respectively. Whereas the proportion of patients who achieved positive anti-SARS-CoV-2 IgG antibody levels after receiving two doses of the vaccine in patients treated with ustekinumab and vedolizumab were 100% and 92.8%, respectively. In patients receiving infliximab and adalimumab, the proportion of patients who had positive anti-SARS-CoV-2 neutralizing antibody levels after two-dose vaccination was 40 out of 59 patients (67.7%) and 14 out 16 patients (87.5%), respectively. Whereas the proportions of patients who had positive anti-SARS-CoV-2 neutralizing antibody levels were 12 out of 13 patients (92.3%) and 13 out of 14 patients (92.8%) in patients receiving ustekinumab and vedolizumab.

**Conclusion:** The majority of patients with IBD on infliximab, adalimumab, and vedolizumab seroconverted after two doses of SARS-CoV-2 vaccination. All patients on ustekinumab seroconverted after two doses of SARS-CoV-2 vaccine. BNT162b2 and ChAdOx1 nCoV-19 SARS-CoV-2 are both likely to be effective after two doses in patients with IBD on biologics. A follow up larger studies are needed to evaluate if decay of antibodies occurs over time.

## Introduction

COVID-19 has rapidly become a major health concern worldwide. It is caused by a new virus called severe acute respiratory syndrome coronavirus 2 (SARS-CoV-2).^1^ To curb the ongoing pandemic caused by SARS-CoV-2 infection, development of vaccines has been advancing at an unprecedented pace, and many countries have authorized different vaccines. BNT162b2 (Pfizer/BioNTech) and the adenovirus-vector vaccine, ChAdOx1 nCoV-19 (Oxford/AstraZeneca) are commonly used worldwide to face further surges of SARS-CoV-2 infection. Both vaccines have demonstrated effectiveness in preventing severe COVID-19 in a phase III placebo-controlled randomized clinical trial and in real-world data.^2–4^

Patients with inflammatory bowel disease (IBD) treated with biologic therapy are considered to be immunocompromised and may be at increased risk of infection.^5,6^ However, in patients with IBD who are receiving biologic therapies, no association was found with severe COVID-19 outcomes such as hospitalization and death.^7^ In addition, the International Organization for the Study of Inflammatory Bowel Disease (IOIBD) recommends that patients with IBD should be vaccinated against COVID-19 and that vaccination should not be deferred in patients receiving biologic therapy.^8^ However, clinical trials excluded patients taking immunosuppressive medications, therefore, it is imperative to evaluate effectiveness of coronavirus disease 2019 (COVID-19) vaccination in patients with IBD with diverse exposure to biologic therapies. Thus, the goal of this study is to determine the immunogenicity of BNT162b2 and ChAdOx1 nCoV-19 vaccines in patients with IBD, receiving different biologic therapies.

## Method

We performed a prospective multi center cohort study at two tertiary care centers (Muabark Alkabeer Hospital and Dasman Center). Patients were recruited at the time of attendance at the gastroenterology infusion rooms and clinics between August 1^st^, 2021, and September 15^th^, 2021. Our inclusion criteria the following: 1) patients with confirmed diagnosis of inflammatory bowel disease before the start of the study 2) patients receiving tumor necrosis factors antagonists (anti-TNFs), vedolizumab or ustekinumab for at least 6 weeks for induction of remission or before next scheduled dose for patients on maintenance therapy (within dose intervals) 3) have received one or two-dose of COVID-19 vaccination with BNT162b2 or ChAdOx1 nCoV-19 vaccine 4) within 4-10 weeks before recruitment for patients who received two doses of vaccination and within 3-6 weeks for patients who received one dose of vaccination 5) at least are 18 years of age or older. Patients were excluded if they were tested positive SARS-CoV-2 previously or had symptoms of COVID-19 since the start of the pandemic. In addition, patients who received corticosteroids two weeks before the first dose of the vaccine up to the time of recruitment were excluded. Also, patients who missed their scheduled biologic dose were excluded. Finally, patients taking immunomodulators such as azathioprine, methotrexate or other immunosuppressive therapies were also excluded. This study was performed and reported in accordance with Strengthening the Reporting of Observational Studies in Epidemiology (STROBE) guideline.^9^

Diagnosis of inflammatory bowel disease (IBD) was made according to the international classification of diseases (ICD-10 version:2016). Patients were considered to have IBD when they had ICD-10 K50, K50.1, K50.8, K50.9 corresponding to Crohn’s disease (CD) and ICD-10 K51, k51.0, k51.2, k51.3, k51.5, k51.8, k51.9 corresponding to ulcerative colitis (UC).^10^

To evaluate the serological response to BNT162b2 or ChAdOx1 nCoV-19 vaccine in patients with IBD on biologic therapies, we measured levels of SARS-CoV-2 Immunoglobulin G (IgG) and neutralizing antibodies post vaccination. In addition, we calculated the percentage of patients who achieved positive SARS-CoV-2 antibodies levels.

We performed descriptive statistics to present the demographic characteristics of patients included in this study and their measured antibody levels. Analysis was conducted using R (R Core Team, 2017). SARS-CoV-2 antibody levels are expressed as medians with interquartile range (IQR) unless otherwise indicated. In addition, percentages of positive IgG and neutralizing antibody levels post vaccination were calculated per each biologic (infliximab, adalimumab, vedolizumab, and ustekinumab).

### Laboratory Methods

In this study, anti-SARS-CoV-2 antibodies quantification from plasma to assess magnitude of systemic virus specific antibodies was performed using enzyme-linked immunosorbent assay (ELISA) kit (SERION ELISA agile SARS-CoV-2 IgG SERION Diagnostics, Würzburg, Germany) based on the manufacturer’s protocol. Units of IgG levels were reported as binding antibody units (BAU)/mL. Values of below 31.5 BAU/mL were considered negative or non-protective. Neutralizing antibody levels below 20% were considered negative or nonprotective. The positive and negative thresholds were determined as per manufacturer’s instructions. Results were construed by calculating inhibition rates for samples as per the following equation: Inhibition = (1 - O.D. value of sample/O.D. value of negative control) ×100%.

### Ethical consideration and role of funders

This study was reviewed and approved by the Ethical Review Board of Mubarak Alkabeer Hospital and Dasman Center “Protocol # RA HM-2021-008” as per the updated guidelines of the Declaration of Helsinki (64th WMA General Assembly, Fortaleza, Brazil, October 2013) and of the US Federal Policy for the Protection of Human Subjects. The study was also approved by the reginal health authority (reference: 3799, protocol number 1729/2021). Subsequently, patient informed written consent was obtained before inclusion in the study.

## Results

### Patients’ characteristics

The study included 126 participants, mean age was 31 years and 60% were males. Mean BMI was 25 kg/m^2^. The proportion of patients with Crohn’s disease and ulcerative colitis was 71% and 29% respectively. (see table 1)

In terms of biologic therapies, 78 (62%) patients were on infliximab and 18 (14%) patients were on adalimumab, whereas the number of patients receiving vedolizumab or ustekinumab was similar at 15 (12%).

**Table 1.**
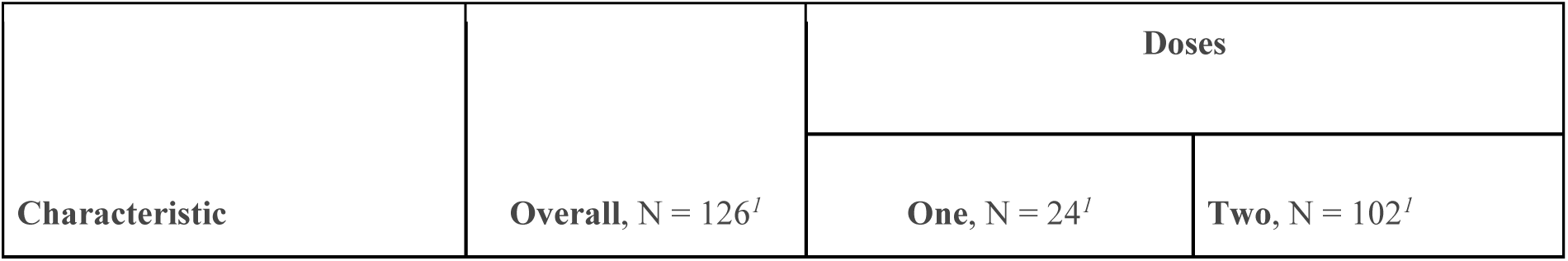
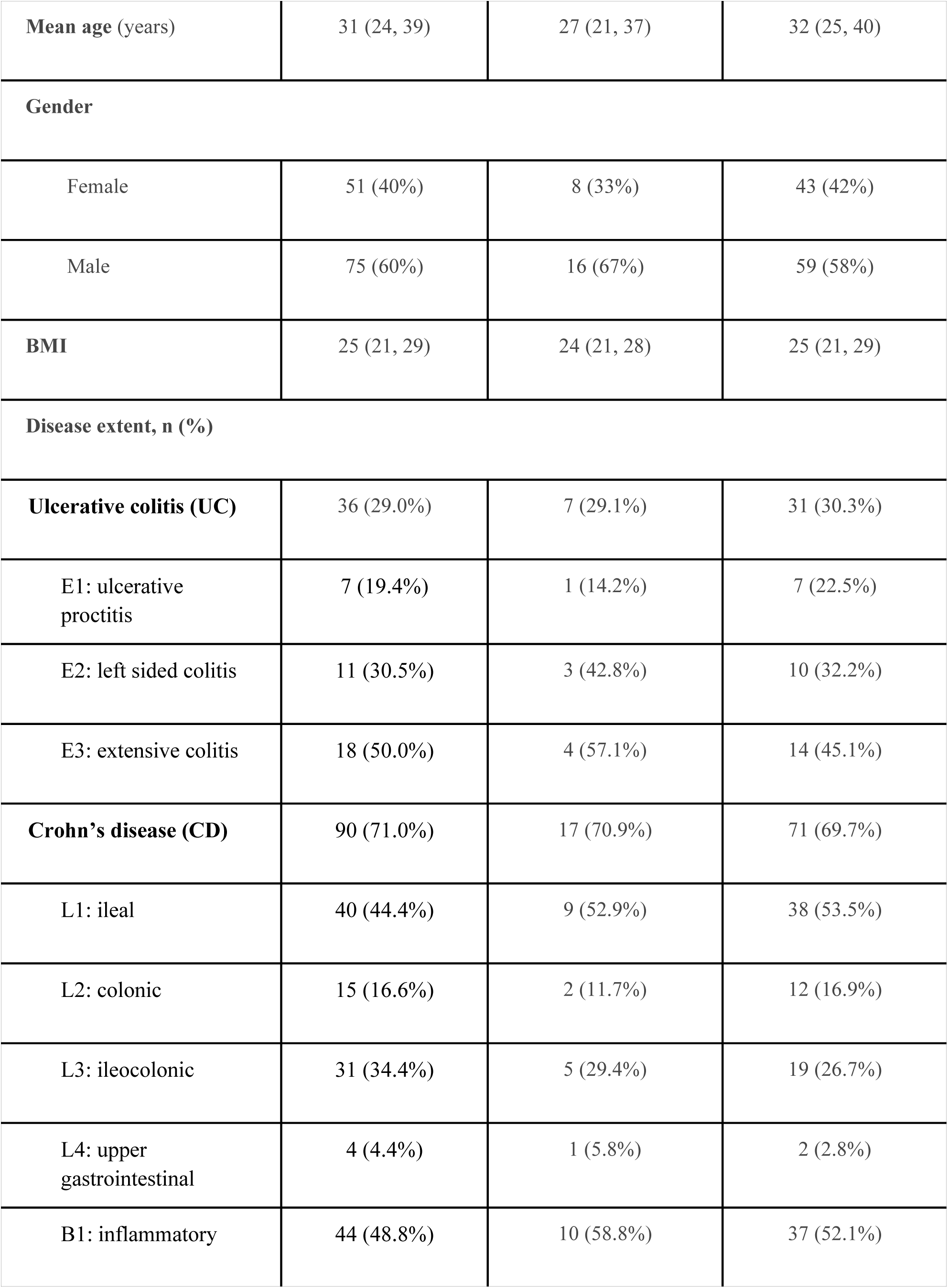
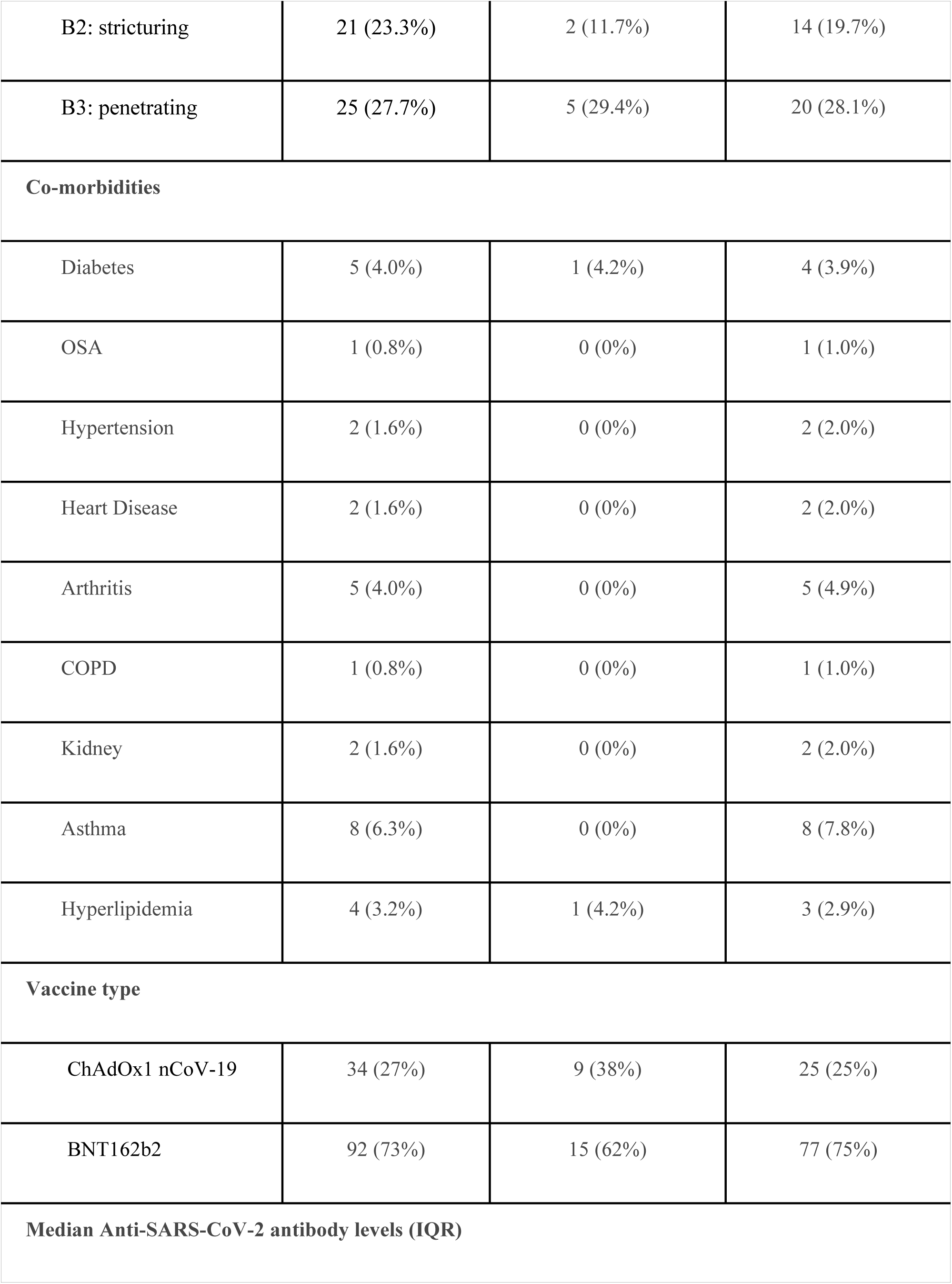
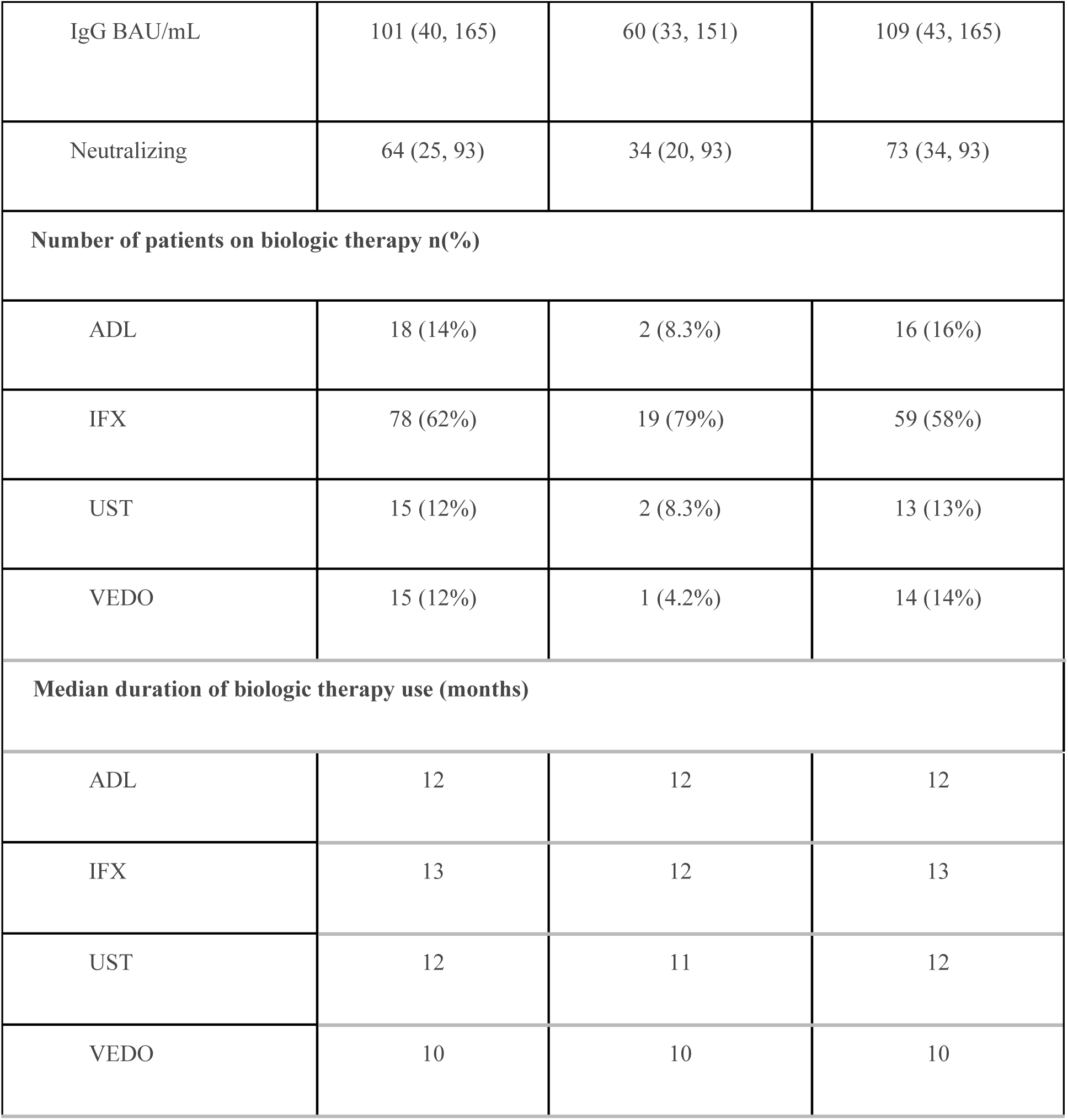
Demographics characteristics of all included patients

| Characteristic | Overall, N = 126<sup>I</sup> | Doses |  |
| --- | --- | --- | --- |
|  |  | One, N = 24<sup>I</sup> | Two, N = 102<sup>I</sup> |
| **Mean age (years)** | 31 (24, 39) | 27 (21, 37) | 32 (25, 40) |
| **Gender** |  |  |  |
| Female | 51 (40%) | 8 (33%) | 43 (42%) |
| Male | 75 (60%) | 16 (67%) | 59 (58%) |
| **BMI** | 25 (21, 29) | 24 (21, 28) | 25 (21, 29) |
| **Disease extent, n (%)** |  |  |  |
| **Ulcerative colitis (UC)** | 36 (29.0%) | 7 (29.1%) | 31 (30.3%) |
| E1: ulcerative proctitis | 7 (19.4%) | 1 (14.2%) | 7 (22.5%) |
| E2: left sided colitis | 11 (30.5%) | 3 (42.8%) | 10 (32.2%) |
| E3: extensive colitis | 18 (50.0%) | 4 (57.1%) | 14 (45.1%) |
| **Crohn's disease (CD)** | 90 (71.0%) | 17 (70.9%) | 71 (69.7%) |
| L1: ileal | 40 (44.4%) | 9 (52.9%) | 38 (53.5%) |
| L2: colonic | 15 (16.6%) | 2 (11.7%) | 12 (16.9%) |
| L3: ileocolonic | 31 (34.4%) | 5 (29.4%) | 19 (26.7%) |
| L4: upper gastrointestinal | 4 (4.4%) | 1 (5.8%) | 2 (2.8%) |
| B1: inflammatory | 44 (48.8%) | 10 (58.8%) | 37 (52.1%) |
| B2: stricturing | 21 (23.3%) | 2 (11.7%) | 14 (19.7%) |
| B3: penetrating | 25 (27.7%) | 5 (29.4%) | 20 (28.1%) |
| **Co-morbidities** |  |  |  |
| Diabetes | 5 (4.0%) | 1 (4.2%) | 4 (3.9%) |
| OSA | 1 (0.8%) | 0 (0%) | 1 (1.0%) |
| Hypertension | 2 (1.6%) | 0 (0%) | 2 (2.0%) |
| Heart Disease | 2 (1.6%) | 0 (0%) | 2 (2.0%) |
| Arthritis | 5 (4.0%) | 0 (0%) | 5 (4.9%) |
| COPD | 1 (0.8%) | 0 (0%) | 1 (1.0%) |
| Kidney | 2 (1.6%) | 0 (0%) | 2 (2.0%) |
| Asthma | 8 (6.3%) | 0 (0%) | 8 (7.8%) |
| Hyperlipidemia | 4 (3.2%) | 1 (4.2%) | 3 (2.9%) |
| **Vaccine type** |  |  |  |
| ChAdOx1 nCoV-19 | 34 (27%) | 9 (38%) | 25 (25%) |
| BNT162b2 | 92 (73%) | 15 (62%) | 77 (75%) |
| **Median Anti-SARS-CoV-2 antibody levels (IQR)** |  |  |  |
| IgG BAU/mL | 101 (40, 165) | 60 (33, 151) | 109 (43, 165) |
| Neutralizing | 64 (25, 93) | 34 (20, 93) | 73 (34, 93) |
| **Number of patients on biologic therapy n(%)** |  |  |  |
| ADL | 18 (14%) | 2 (8.3%) | 16 (16%) |
| IFX | 78 (62%) | 19 (79%) | 59 (58%) |
| UST | 15 (12%) | 2 (8.3%) | 13 (13%) |
| VEDO | 15 (12%) | 1 (4.2%) | 14 (14%) |
| **Median duration of biologic therapy use (months)** |  |  |  |
| ADL | 12 | 12 | 12 |
| IFX | 13 | 12 | 13 |
| UST | 12 | 11 | 12 |
| VEDO | 10 | 10 | 10 |

92 patients were vaccinated with BNT162b2 (73%) and 34 patients were vaccinated with ChAdOx1 nCoV-19 (27%). 102 patients received two doses of SARS-CoV-2 vaccine, BNT162b2 or ChAdOx1 nCoV-19, whereas 24 patients received only one dose.

### Serological Response to both BNT162b2 and ChAdOx1 nCoV-19 Vaccines

The proportion of patients who achieved positive anti-SARS-CoV-2 IgG antibody levels after receiving 2 doses of either BNT162b2 or ChAdOx1 nCoV-19 vaccines in patients treated with infliximab and adalimumab were 44 out of 59 patients (74.5%) and 13 out of 16 patients (81.2%), respectively. Whereas the proportion of patients who achieved positive anti-SARS-CoV-2 IgG antibody levels after receiving two doses of the vaccine in patients treated with ustekinumab and vedolizumab were 100% and 92.8%, respectively. In patients receiving infliximab and adalimumab, the proportion of patients who had positive anti-SARS-CoV-2 neutralizing antibody levels after two-dose vaccination was 41 out of 59 patients (69.4%) and 14 out 16 patients (87.5%), respectively. Whereas the proportions of patients who had positive anti-SARS-CoV-2 neutralizing antibody levels were 12 out of 13 patients (92.3%) and 13 out of 14 patients (92.8%) in patients receiving ustekinumab and vedolizumab.

### Serological Response to BNT162b2 vaccine

In patients vaccinated with BNT162b2 and receiving infliximab, the proportion of participants with positive anti-SARS-CoV-2 IgG levels was 34 out of 43 patients (79%), whereas the proportion of patients treated with adalimumab and had positive SARS-CoV-2 IgG levels was 12 out of 15 patients (80%). The proportion of patients treated with infliximab and adalimumab who had positive anti-SARS-CoV-2 neutralizing antibody levels was 30 out of 43 (69.7%) and 13 out of 15 patients (86.6%), respectively. Finally, the proportion of patients treated with ustekinumab and vedolizumab who had positive anti-SARS-CoV-2 IgG levels was 100% and 90.9%, respectively.

### Serological Response to ChAdOx1 nCoV-19 vaccine

In patients vaccinated with ChAdOx1 nCoV-19 vaccine receiving infliximab, the proportion of participants with positive anti-SARS-CoV-2 IgG and neutralizing antibody levels was 10 out of 16 (62.5%). The proportion of patients treated with ustekinumab and vedolizumab who had positive anti-SARS-CoV-2 IgG and neutralizing antibody levels was 100%. Only one patient on adalimumab received ChAdOx1 nCoV-19 vaccine. The levels of anti-SARS-CoV-2 IgG, and neutralizing antibody was above the positive threshold for that patient.

### Serological Response to one dose of either BNT162b2 or ChAdOx1 nCoV-19 vaccine

In total, 24 patients of our cohort received a single dose of BNT162b2 or ChAdOx1 nCoV-19 vaccines. 18 (75%) patients had positive anti-SARS-CoV-2 IgG and neutralizing antibody levels 3-6 weeks after first dose. Out of the total patients who received a single dose of the vaccine, 19 were on infliximab, 2 patients were on adalimumab, 2 were on ustekinumab, and 1 was on vedolizumab. In patients receiving infliximab, 14 out of 19 (73.6%) had positive anti-SARS-CoV-2 IgG and neutralizing antibody levels.

### Median SARS-CoV-2 IgG and Neutralizing antibody levels

In patients who received two doses of the BNT162b2 vaccine and were treated with infliximab, the median [interquartile range (IQR)] anti-SARS-CoV-2 IgG level is 103 BAU/mL (38, 181), whereas the median was 90 BAU/mL (48, 134) in patients receiving adalimumab. Respective anti-SARS-CoV-2 IgG levels were 130 BAU/mL (104, 159) and 139 BAU/mL (117, 180) in patients treated with ustekinumab and vedolizumab. (figure 1) Neutralizing antibody levels were 76 (17, 95) and 59 (32, 71) in patients treated with infliximab and adalimumab respectively. Neutralizing antibody levels in patients ustekinumab and vedolizumab users were 84 (69, 88) and 88 (79, 95) respectively. (See table 2)

**Figure 1.**
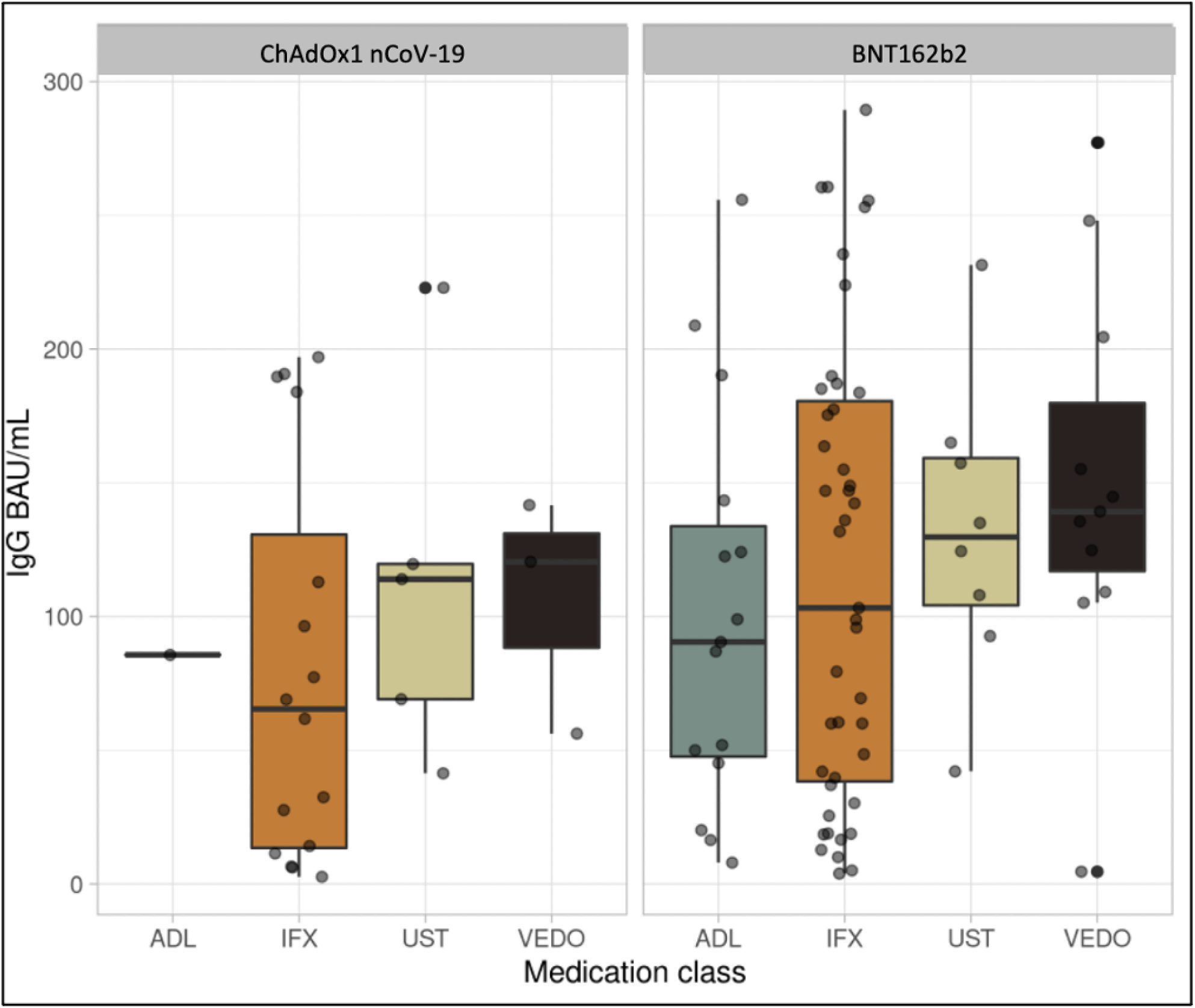
Box plot illustrating IgG antibody concentrations among patients on different biologic therapies.

**Table 2:** Serological response by Biologics in participants vaccinated with BNT162b2 vaccine

| Characteristic | Overall, N = 77<sup>1</sup> | ADL, N = 15<sup>1</sup> | IFX, N = 43<sup>1</sup> | UST, N = 8<sup>1</sup> | VEDO, N = 11<sup>1</sup> |
| --- | --- | --- | --- | --- | --- |
| **IgG BAU/mL** | 124 (49, 175) | 90 (48, 134) | 103 (38, 181) | 130 (104, 159) | 139 (117, 180) |
| **Neutralizing** | 76 (34, 94) | 59 (32, 71) | 76 (17, 95) | 84 (69, 88) | 88 (79, 95) |
| <sup>1</sup> Median (IQR) or Frequency (%) |  |  |  |  |  |

In patients who received two doses of the ChAdOx1 nCoV-19 vaccine and were treated with infliximab, the median (IQR) anti-SARS-CoV-2 IgG level was 65 BAU/mL (14, 131). Respective anti-SARS-anti-SARS-CoV-2 IgG levels were 114 BAU/mL (69, 120) and 120 BAU/mL (88, 131) in patients treated with ustekinumab and vedolizumab. Median neutralizing antibody levels were 49 (10, 84) in patients treated with infliximab. (Figure 2) Median neutralizing antibody levels in patients ustekinumab and vedolizumab users were 82 (37, 86) and 61 (42, 75) respectively. (See table 3) Finally, in patients who received one dose, median (IQR) anti-SARS-CoV-2 IgG and neutralizing antibody levels were 60 (33, 151) and 34 (20, 93), respectively. (See table 4)

**Figure 2.**
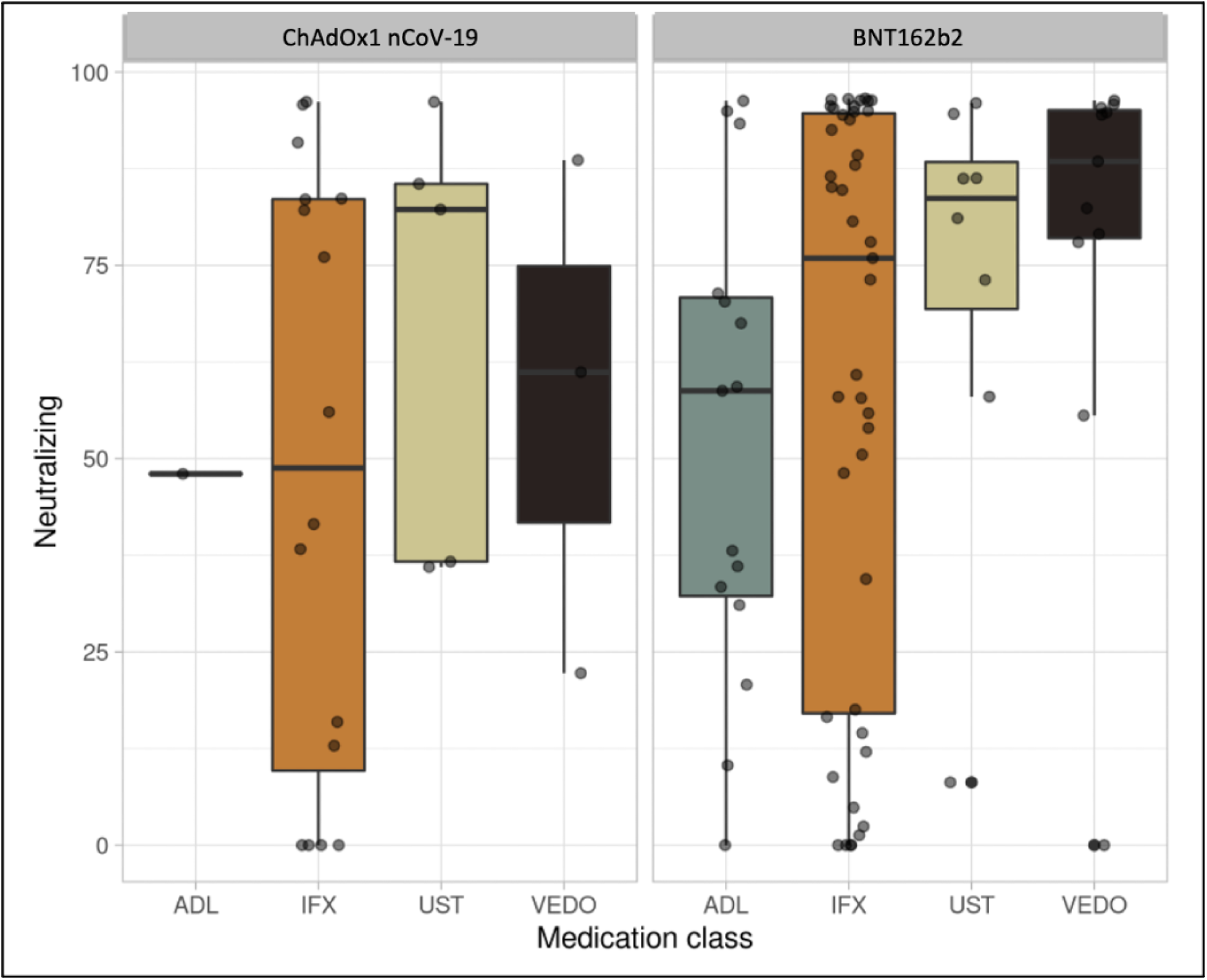
Box plot illustrating Neutralizing antibody concentrations among patients on different biologic therapies.

**Table 3:** Serological response by Biologics in participants vaccinated with or ChAdOx1 nCoV-19

| Characteristic | Overall, N = 25<sup>1</sup> | ADL, N = 1<sup>1</sup> | IFX, N = 16<sup>1</sup> | UST, N = 5<sup>1</sup> | VEDO, N = 3<sup>1</sup> |
| --- | --- | --- | --- | --- | --- |
| **IgG BAU/mL** | 77 (32, 120) | 86 (86, 86) | 65 (14, 131) | 114 (69, 120) | 120 (88, 131) |
| **Neutralizing** | 56 (22, 84) | 48 (48, 48) | 49 (10, 84) | 82 (37, 86) | 61 (42, 75) |
| <sup>1</sup> Median (IQR) or Frequency (%) |  |  |  |  |  |

**Table 4:**
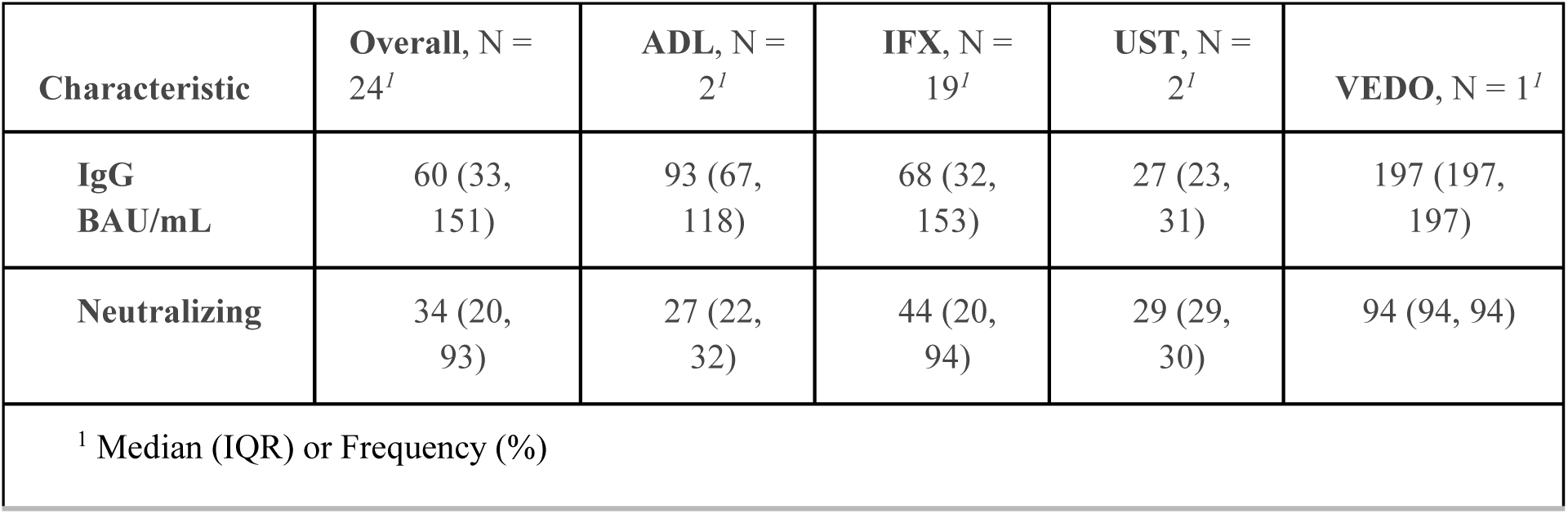
Serological response among patients who has taken only one dose of either BNT162b2 or ChAdOx1 nCoV-19 vaccine

## Discussion

In this study we evaluated the serological response to one or two doses of BNT162b2 and ChAdOx1 nCoV-19 vaccines in a cohort of 126 patients with IBD and receiving different biologic therapies. Most patients seroconverted, had positive antibody responses, 4-10 weeks after the second vaccine dose. Interestingly, while the majority of patients (70-90%) on anti-TNFs or vedolizumab achieved positive anti-SARS-CoV-2 IgG and neutralizing antibody levels, all patients receiving ustekinumab achieved positive anti-SARS-CoV-2 antibody levels regardless of the vaccine used.

Our findings reinforce the findings of previous studies indicating positive humoral immune response with complete vaccination in patients with IBD treated biologic therapies. Deepak et al found that in a cohort of 133 participants with different chronic inflammatory conditions and receiving immunosuppressive therapy, 88.7% had detectable anti-SARS-CoV-2 antibody levels after 2 doses of the BNT162b2 or mRNA-1273 vaccine. However, the mean concentration of anti-SARS-CoV-2 antibodies in patients with inflammatory disease was lower compared to immunocompetent participants.^11^

Our findings showing seroconversion across medication groups are consistent with other IBD studies. One study^12^ assessed humoral responses after mRNA vaccination in adults with IBD at different time points. The study found that regardless of medication regimen, 99% of participants had detectable antibodies after 2 weeks. In addition, anti-SARS-CoV-2 IgG levels were lowest in patients receiving anti–tumor necrosis factor combination therapy or corticosteroids. However, it is important to mention that the study was not powered to assess differences between medication subgroups.

Khan et al explored the effectiveness of mRNA vaccination in the veterans patients with IBD receiving diverse immunosuppressive medications.^13^ The authors observed that two doses of mRNA vaccine, but not a single dose, was associated with 69% reduced hazard of infection relative to an unvaccinated status. They also suggested that SARS-CoV-2 vaccine effectiveness is similar in patients with IBD regardless of the immunosuppressive agents being used. Wong et al reported serological responses to mRNA vaccination in patients with IBD on biologic therapies.^14^ They found that despite achieving antibody levels consistent with presumed protection, there was an association of lower antibody levels in patients with vedolizumab for all antibodies tested and with anti-TNFs for the receptor binding domain (RBD) of the SARS-CoV-2 S protein total immunoglobulin.

One study^15^ reported the effectiveness of the BNT162b2 mRNA COVID-19 vaccine in patients with IBD receiving different therapies including biologic. The authors found that in patients with IBD, the vaccine was highly effective and comparable to the general population with an absolute breakthrough infection rate of 0.1% for fully vaccinate IBD patients. However, the study suggested that patients with Crohn’s disease (CD) may have an increased risk for breakthrough infection, but larger studies are needed to validate this finding.

Anti-SARS-CoV-2 circulating antibodies play a major role in risk reduction of severe COVID-19. IgG class particularly target the spike protein of SARS-CoV-2, its S1 subunit or its receptor binding domain (RBD).^16^ Thus, the binding of the virus to host receptors is weakened or terminated completely. Neutralizing antibodies are major contributors to immunity.^17,18^ Vaccine studies showed that high levels of IgG and neutralizing antibodies against the SARS-CoV-2 spike protein correlate with protection. However, it is not currently known what the threshold titers should be to achieve protection.

CLARITY IBD study^19^ showed that the lowest rates of seroconversion were observed in participants treated with infliximab in combination with an immunomodulator with both the BNT162b2 or ChAdOx1 nCoV-19 vaccines. Interestingly, highest rates of seroconversion were seen in patients treated with vedolizumab monotherapy who received either one of the previously mentioned vaccines.

Anti-TNF agents inactivate the proinflammatory cytokine TNF by direct neutralization, thus resulting in suppression of inflammation. The cytokine TNF is involved in multiple aspects of host immune responses, including T-cell dependent antibody production. This mechanism of action may in part explain why TNF blockade is clinically beneficial, but also explain the increased risk of serious and opportunistic infections and impaired response to some vaccines.^20^ The mechanism of action of vedolizumab, an anti-integrin, is hypothesized to be restricted to the gastrointestinal tract, not affecting immune responses outside of the gastrointestinal tract.^21^ Finally, ustekinumab inhibit the p40 subunit of interleukin (IL)-12/23, which are major drivers of the adaptive immune response. ^22^

Taken together, these emerging data provide reassurance that biologic therapies do not markedly reduce the response to COVID-19 immunization and support recent consensus recommendations to vaccinate all patients with IBD regardless of immune-modifying therapies.^23^ However, patients should be advised that vaccine efficacy may be decreased when receiving systemic corticosteroids. In addition, anti-SARS-CoV2 antibodies decay remains a legitimate concern in patients with IBD on biologic therapy,^24^ therefore, booster doses are being recommended by official organizations.^25,26^

Our study has several strengths. It is a multi-center prospective study. It examined the effect of two important SARS-CoV-2 vaccines in a vulnerable patient population with IBD on common biologic agents, who might be reluctant to vaccinate. It is well designed with low risk of bias given its inclusion and exclusion criteria. It addresses a very clinically relevant question that physicians and patients are eager to know.

However, our study has some limitations. It includes a small sample size of patients, so it was not powered to assess differences across medication subgroups. Furthermore, only humoral serological responses were assessed, it is likely that T cell-mediated immunity plays a possible protective role. Finally, given the observational nature of the study, possible confounders may exist, such as difference in biologic dosing and serum drug concentrations were not accounted for. A follow up larger studies needed to evaluate if decay of antibodies occurs over time and if a possible third booster vaccine dose may be needed.

To conclude, this study showed that most patients with IBD on infliximab, adalimumab, and vedolizumab seroconverted after two doses of SARS-CoV-2 vaccination. All patients on ustekinumab seroconverted after two doses of SARS-CoV-2 vaccine. BNT162b2 and ChAdOx1 nCoV-19 SARS-CoV-2 are both likely to be effective after two doses in patients with IBD on biologics. A follow up larger studies are needed to evaluate if decay of antibodies occurs over time and if a third booster vaccine dose may be needed.

## Supporting information

STROBE checklist

## Data Availability

All data produced in the present study are available upon reasonable request to the authors

## Acknowledgement

none

